# Genetic determinants of interventricular septal anatomy and the risk of ventricular septal defects and hypertrophic cardiomyopathy

**DOI:** 10.1101/2021.04.19.21255650

**Authors:** Mengyao Yu, Andrew R. Harper, Matthew Aguirre, Maureen Pittman, Catherine Tcheandjieu, Dulguun Amgalan, Christopher Grace, Anuj Goel, Martin Farrall, Ke Xiao, Jesse Engreitz, Katherine Pollard, Hugh Watkins, James R. Priest

## Abstract

**Background:** The interventricular septum (IVS) plays a primary role in cardiovascular physiology and a large proportion of genetic risk remains unexplained for structural heart disease involving the IVS such as hypertrophic cardiomyopathy (HCM) and ventricular septal defects (VSD).

**Objectives:** We sought to develop a reproducible proxy of IVS structure from standard medical imaging, discover novel genetic determinants of IVS structure, and relate these loci to two rare diseases of the IVS.

**Methods:** We performed machine learning to estimate the cross-sectional area of the interventricular septum (IVS.csad) obtained from the 4-chamber view of cardiac MRI in 32,219 individuals from the UK Biobank. Using these extracted measurement of IVS.csad we performed phenome-wide association to relate this proxy measure to relevant clinical phenotypes, followed by genome-wide association studies and Mendelian Randomization.

**Results:** Automated measures of IVS.csad were highly accurate, and strongly correlated with anthropometric measures, blood pressure, and diagnostic codes related to cardiovascular physiology. A Single nucleotide polymorphism in the intron of *CDKN1A* was associated with IVS.csad (rs2376620, Beta 8.4 mm2, 95% confidence intervals (CI) 5.8 to 11.0, p=2.0e-10), and a common inversion incorporating *KANSL1* predicted to disrupt local chromatin structure was associated with an increase in IVS.csad (Beta 8.6 mm2, 95% CI 6.3-10.9, p=1.3e-13). Mendelian Randomization suggested that inheritance of a larger IVS.csad was causal for HCM (Beta 2.45 log odds ratio (OR) HCM per increase in SD of IVS.csad, standard error (SE) 0.48, pIVW = 2.8e-7) while inheritance of a smaller IVS.csad was causal for VSD (Beta −2.06 log odds ratio (OR) VSD per decrease in SD of IVS.csad, SE 0.75, pIVW = 0.006)

**Conclusion:** Automated derivation of the cross sectional area of the IVS from the 4-chamber view allowed discovery of loci mapping to genes related to cardiac development and Mendelian disease. Inheritance of a genetic liability for either large or small interventricular septum, appears to confer risk for HCM or VSD respectively, which suggests that a considerable proportion of risk for structural and congenital heart disease may be localized to the common genetic determinants of cardiovascular anatomy.

## INTRODUCTION

Within the cardiac ventricles, the interventricular septum (IVS) separates deoxygenated pulmonary blood flow from oxygenated systemic blood flow and is a foundation of mammalian physiology^1^. The IVS is the site of two important diseases; ventricular septal defects (VSD) are among the most common forms of congenital heart disease, where a thin or abnormally developed IVS leaves a communication linking the lumens of the left and right ventricle which when untreated may lead to heart failure and pulmonary hypertension^2^. Alternatively, the increased thickness and altered geometry of the IVS in hypertrophic cardiomyopathy (HCM) may progress to the point of obstruction of the left-ventricular outflow tract.^3^ While both VSD and HCM are primarily genetic in origin, Mendelian inheritance does not fully account for the risk of HCM^4^ or for individual variability in disease amongst individuals carrying the same pathogenic variant. Monogenic disease causing VSD is very rare, and the genetic basis of VSD is simply not known in the majority of affected individuals^5^.

The IVS has two primary developmental origins, with the membranous portion arising from mesenchymal lineages in the cardiac cushions^6^ and the muscular portion developing from ingrowth of the primary heart tube^7^. A complex microanatomical orientation of cardiomyocyte fibers through the IVS links the function of the right and left ventricles^8^. After adaptation to postnatal circulation over the first months of life, the mature IVS has a complex relationship with normal physiology, with the thickness of the IVS increased in both exercise capacity^9^ as well as in hypertension^10^.

Here we derive a simple automated measure of the IVS derived from MRI imaging of the heart in the 4-chamber view, IVS cross-sectional area at diastole (IVS.csad). We show that IVS.csad is correlated with standard clinical measures and is useful as suitable proxy for the architechture of the IVS. We report genetic correlates of IVS.csad in 32,219 individuals derived from cardiac MRI of the UK Biobank. Using Mendelian Randomization, we describe new causal relationships linking smaller IVS.csad with risk for VSD and larger IVS.csad with risk for HCM.

## METHODS

### Ethics statement

The NHS National Research Ethics Service (ref: 11/NW/0382) granted ethical approval for the distribution of deidentified imaging, genetic, and medical record data from the UK Biobank (UKB) for any qualified researcher.

### UK Biobank cohort and IVS cross sectional area measurement from cardiac MRI

We built a U-Net segmentation model of cine images of the 4-chamber (4Ch) view of the heart with pre-trained weights from VGG11, which was further trained on 60 hand-labeled 4Ch images, validated on 20 hand-labeled images, and tested on 20 hand-labeled images^11^. This approach yielded a validation dice score of 91.2% and a test dice score of 93.8% which were equivalent to differences observed between expert human annotators in an open-source deep learning framework upon which this work is based^12^. We then applied the segmentation model to 32,219 4Ch images from the UKB and generated masks for all 4 chambers including the left and right ventricles and the left and right atrium. Following generation of the masks from the trained U-Net segmentation model, we used another function to measure the interventricular septal areas. We first located the base of the atriums and apex of the ventricles along the medial axis of the chambers, and secondly we located the annuli of the atrioventricular valves using the intersecting lines of the atriums and the ventricles, then finally we use the contour lines of the masks of atriums and ventricles between the annulus lines and the base of the atriums or the apex of the ventricles to enclose the area of the interventricular septum. The metadata in the dicom measurements were used to convert from pixels^2^ to mm^2^. The frame representing end-diastole was obtained by selecting the image frame with the largest estimate of left ventricular volume provided by the U-Net segmentation algorithm.

To exclude outliers related to imaging error or methodological inaccuracy, automated measurements were plotted relative to body surface area with standard measures of quality control. We excluded measures +/-3 standard deviations. Manual annotation of 50 randomly selected images spanning the cardiac cycle was performed by a clinician (JRP) blinded to automated measures or anthropometric characteristics. The percent difference between automated and manual measurements ([automated measure - manual measure]/manual measure) was examined for systematic relationships to anthropomorphic predictors (Age in years, body surface area, genetic sex). Body surface area was estimated from height and weight using the Haycock formula^13^.

### Phenome wide association study

Given that IVS.csad is not a standard clinical measurement, using previously described methods^14^ we performed a phenome-wide association studies (PheWAS) to highlight clinical associations with 67 cardiovascular phenotypes aggregated from ICD-10 codes as phecodes^15^. For the reporting of PheWAS results, we excluded phenotypes with less than 50 individuals (for continuous traits) or less than 50 cases (for binary-coded traits), and controled the positive false discovery rate (pFDR). We performed PheWAS for the IVS.csad amongst individuals with MRI data (n=32,219).

### GWAS, Burden testing, and Annotation

The UK Biobank data release available at the time of analysis included genotypes for 488,377 participants, obtained through either the custom UK Biobank Axiom array or the Affymetrix Axiom Array. Genotypes were imputed to the TOPMed panel (Freeze5) at the Michigan imputation server. Only variants with minor allele frequency (MAF) greater than or equal to 0.01, and minor allele count (MAC) greater or equal to 5, and variants which have Hardy-Weinberg equilibrium exact test p-value greater than 1e-20 in the entire MRI dataset and an empirical-theoretical variance ratio (MaCH r^2^) threshold above 0.3 were included. The main GWAS was conducted on the largest subset of participants with MRI data from the largest unrelated European-ancestry cohort defined using the variable in.white.British.ancestry.subset in the file ukb_sqc_v2.txt provided as part of the UKB data release (n = ∼27,100 individuals with estimates derived from imaging data, age = 55.0±7.4). To replicate the findings, we separated the dataset into a discovery set of 22,124 participants and a replication set of 4,899 participants released at a later date. To further explore the findings from the European GWAS, we included three independent sets of other ethnic backgrounds with cardiac MRI who were not included in the discovery set, including African/Afro-Caribbean (AF, n = 222, age = 49.6±7.0), East Asian (EAS, n = 85, age = 49.2±5.5), and South Asian (SAS, n = 368, age = 52.1±7.9). Examination of those samples according the genetic principal components showed that many were mostly of non-European ancestry and were unrelated [Table S1]. Testing of single nucleotide variants and indels was performed using an using linear regression PLINK^16^2 (v2.00a2LM) additive model^17,18^, including gender, age, BSA, and genetic principal components 1-4. For genic and regional CNV burden tests, associations were performed using linear regression PLINK2 (v2.00a2LM) additive model^17,18^, including gender, age, BSA, Principal components 1-10 and the length and total number of CNV per individual as covariates as previously described^19^. Locuszoom or custom python scripts were used to generate regional association plots^20^. Trans-ethnic meta-analysis was performed using the METAL software using the standard error analysis scheme.

Independent SNPs were identified by linkage disequilibrium (LD) r2 0.6 and were annotated to cardiac eQTL (GTEx v8: heart atrial appendage and heart left ventricle), CADD, RDB, and the GWAS catalog. GWAS of rare variant and gene-based tests were performed using score and SKATO as implemented in the RVTEST package (link). For the RV-GWAS, age, sex, BSA, Principal components 1-10 were used as covariates to calculate the association of 6 million (6,593,945) imputed/genotyped variants of IVS with minor allele frequency between 0.01 and 0.0005, minor allele count greater than 15, an imputation quality MachR2 greater than 0.8, and p-value for Hardy Weinberg Equilibrium greater than 1e-20. For gene-based tests, the selected variants were mapped to the UCSC GRCh38 refGene annotations, where every unique protein-coding gene (n = 22,933) listed was tested.

### In silico analyses for contact prediction and enhancer activity

To predict changes in 3D chromatin folding brought about by the chr17 inversion, we used Akita^21^, a convolutional neural network model trained to predict Hi-C maps from about one megabase of DNA. We first validated that Akita can reproduce wildtype contact frequencies in the region of [GRCh38/hg38] chr17:45392804-46440468. We then used the inversion sequence as input to generate a mutant Hi-C prediction. Comparing this to the wildtype, we predicted changes in contact frequency between promoters, gene bodies, and candidate enhancer elements in the GeneHancer database^22^. To determine possible regulatory functions for single variants in GWAS loci, we examined all variants with P < 10-6. We intersected these variants with predicted enhancers in 131 cell types identified by the activity-by-contact (ABC) Model, which combines measurements of enhancer activity (based on ATAC-seq, DNase-seq, and H3K27ac ChIP-seq) with estimates of enhancer-promoter 3D contact frequencies (based on Hi-C)^23,24^. We examined DNase-seq and ATAC-seq data across a range of cardiovascular cell types from the ENCODE Project. Finally, we examined sequence motif predictions for identified variants using a database of transcription factor binding site motifs^25^.

### LDSC and genetic correlation analyses

To calculate genetic correlation between polygenic risk score of IVS cross sectional area annular size and other related phenotypes, we obtained summary statistics for cardiac MRI-derived LV measurements (left ventricular end-diastolic volume (LVEDV), left ventricular end-systolic volume (LVESV), stroke volume (SV), the body-surface-area (BSA) indexed versions for cardiovascular traits (LVEDVi, LVESVi, and SVi), and left ventricular ejection fraction (LVEF)^26^, atrial fibrillation (AF)^27^, nonischemic cardiomyopathy (NICM)^28^, heart failure^29^, heart failure using UK Biobank data^28^, hypertension^30^, PR Interval^31^, Myocardial Infarction (MI) and coronary artery disease (CAD)^32^, heart rate^33^, and IVS cross sectional area prolapse (MVP)^34^. Using these data we performed LD Score regression^35^ based on the reformatted summary statistics filtered to HapMap3.

### Mendelian Randomization

We performed two-sample Mendelian randomization (MR) to test for causal relationships between IVS.csad and risk of two structural diseases of the IVS, specifically VSD and HCM. Summary statistics from a VSD GWAS that considered 191 individuals with VSD and 5,159 controls were retrieved.^26^ Summary statistics from a multi-ancestry HCM GWAS that evaluated 2,780 HCM cases and 47,486 controls were retrieved^36^. An instrumental variable (IV), consisting of 22 variants was generated by first selecting variants with p-value < 5e-06 from a standardized trans-ethnic meta-analysis of IVS.csad and then, applying a linkage disequilibrium clumping procedure (r^2^ of 0.01 across a window of 1000 kb) via TwoSampleMR (https://github.com/MRCIEU/TwoSampleMR/). The F statistic for this 22 SNP instrument was 15.1 and accounts for 1.04% SNP heritability.

We performed two-sample MR using four different methods, specifically inverse-variance weighted (IVW), weighted median, MR-Egger, and MR-PRESSO (mendelian randomisation pleiotropy residual sum and outlier) using MR-Base^37,38^. Each method assumes a different set of underlying assumptions; all variants included in the instrument are assumed to be valid in the IVW method, and a fixed effects IVW model assumes each variant confers the same mean effect, with no horizontal pleiotropy. A random effects IVW model, accounts for the possibility that variants included in the instrument yield different mean effects and can provide an unbiased estimate when horizontal pleiotropy is balanced^39^. The MR-Egger regression estimates the relationship of effects across instruments functioning as a sensitivity analysis for directional pleiotropy^40^, weighted median offers a consistent estimate of effect size when a minimum of 50% of the weights come from valid IVs, and the MR-PRESSO model measures and robustly accounts for the presence of horizontal pleiotropy^38^.

## RESULTS

After exclusion of outliers we obtained automated estimates of the cross-sectional area of the interventricular septum during ventricular diastole (IVS.csad) for 31,587 individuals yielding a mean IVS.csad of 651 mm^2^ (range 207 – 1108 mm^2^, standard deviation 166 mm^2^) [Fig.1A]. In comparison to blinded manual measurements, automated measures were on average 24 mm^2^ or 3.2% smaller than manual measures of IVS.csad with no systematic relationship of measurement error to body surface area (BSA), genetic sex, or age across 50 randomly selected test images [Table S2].

Given that IVS.csad is not a standard clinical measure in MRI or echocardiography and is infrequently characterized in the literature^27^ we sought to provide clinical context. Similar to the size of other cardiac structures, IVS.csad scales with BSA in a linear fashion [Fig.1B]. Amongst the 31,587 individuals with an automated estimate, IVS.csad was also correlated with genetic sex and body mass index (BMI) (Pearson’s r > 0.6), moderately correlated with systolic and diastolic blood pressure (Pearson’s r > 0.3), and mildly correlated with birthweight (Pearson’s r > 0.1) [Fig.1C]. Additionally, in a PheWAS of IVS.csad for 67 cardiovascular phenotypes we observed that a larger IVS.csad was positively associated with PheCODES encompassing “abnormal heart sounds” (p_fdr_ = 0.003, Odds Ratio (OR) 2.1 per standard deviation (SD) increase in IVS.csad, ICD10 codes R00 - R01.3), “non-rheumatic aortic valve disease” (p_fdr_ = 0.003, OR 2.4 per SD IVS.csad, ICD10 I08.2, I35, I35.9), and “abnormal functional study of the cardiovascular system” (p_fdr_ = 0.02, OR 2.6 per SD IVS.csad, ICD10 R94.3[Fig.1D]. Importantly, in a random subset of 50 individuals selected for manual measurement, IVS.csad was correlated with maximal interventricular septal thickness (Pearson’s r = 0.62), a standard clinical measure of IVS structure^28^.

**Figure 1.**
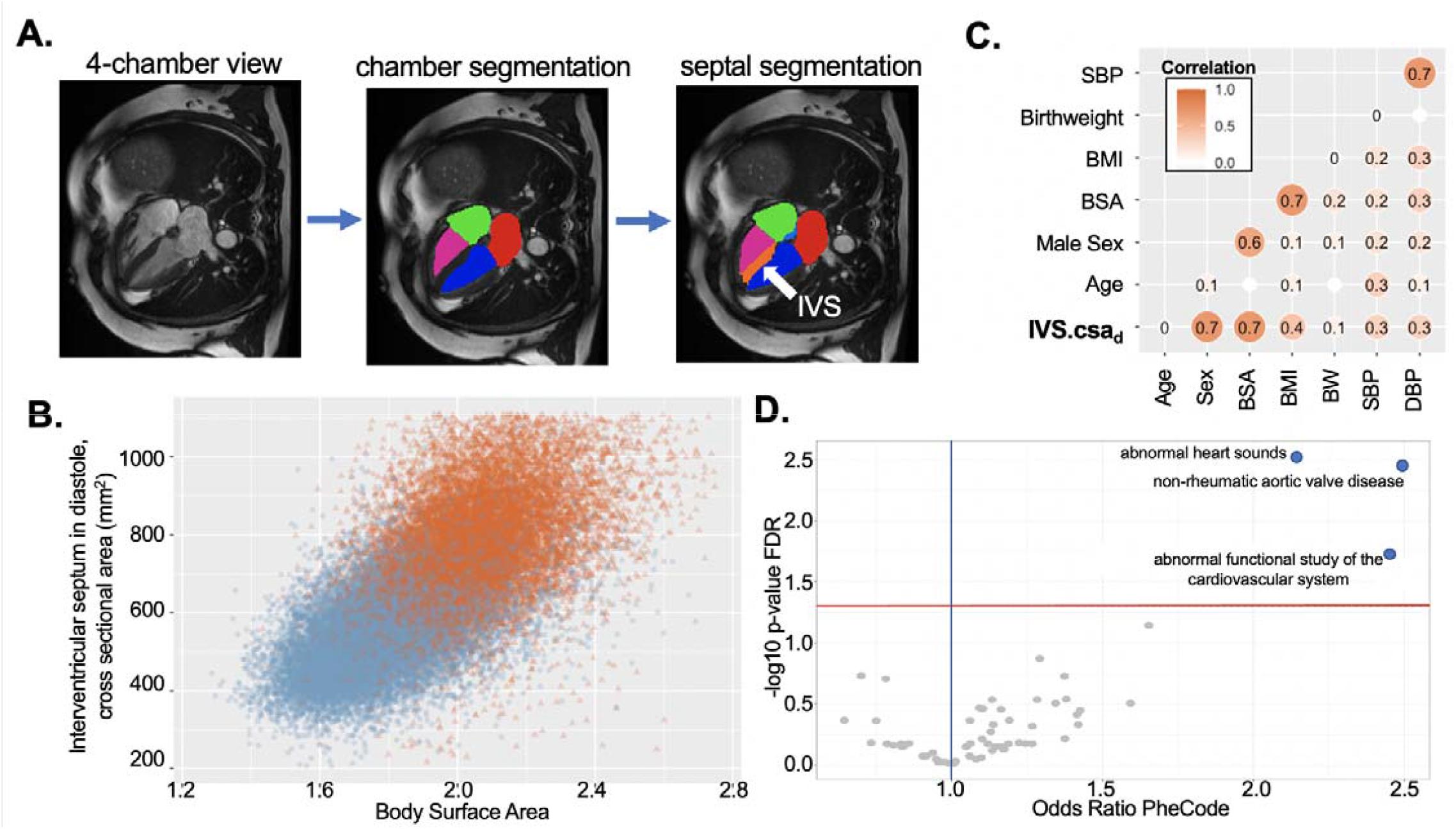
Cross-sectional area in diastole is a new proxy for structure and function of the interventricular septum. **A**. Schema of cardiovascular measurements. Still frame of a cardiac MRI in the four-chamber view (left panel) demonstrating segmentation of lumen of the cardiac chambers (middle panel) with the left ventricle indicated by the blue region, left atrium indicated by the red region, right atrium indicated by the green region, and right ventricle in magenta. Chamber segmentation is followed by segmentation of the interventricular septum (orange, indicated by white arrow). **B**. The distribution of interventricular septal cross-sectional area in diastole (IVS.csad). Values are plotted in mm^2^ relative to body surface area (BSA), orange and blue points represent male and female participants respectively. In a simple linear model including Sex, Age, systolic and diastolic blood pressure and body mass index, BSA explains 44.8% of the observed variation in IVS.csad (284 mm^2^ IVS.csad per m^2^ BSA, p < 2e-16). **C**. Correlation plot of IVS.csad versus a standard set of clinical measures. IVS.csad is most strongly correlated with genetic sex and BSA, moderately correlated with BMI and blood pressure, and slightly correlated with birthweight. Notably there was no strong correlation with age. All non-zero Pearson’s correlations reported are strongly statistically significant (p < 5e-08). **D**. Volcano plot representing the PheWAS of IVS.csad for 67 cardiovascular phenotypes grouped as PheCODES. The X-axis represents the association with the PheCODE per standard deviation change in the IVS.csad, and the absolute value of Y-axis represents the negative logarithm of the p-value adjusted with a standard false discovery rate. Across an unbiased assessment of clinical phenotypes, increased IVS.csad was positively associated with the clinical findings of abnormal heart sounds, non-rheumatic aortic valve disease, and abnormal functional studies of the cardiovascular system.

Given that IVS.csad was strongly correlated with key clinical measures of cardiovascular structure and function, we performed a GWAS of IVS.csad as a proxy for interventricular septal mass in 26,844 individuals of European ancestry divided into discovery (n = 21,945) and replication (n = 4,899) subsets [Table S1]. Using the summary statistics, we performed LD-score correlation (LDSC) to relate the genetic determinants of IVS.csad to a set of common cardiovascular traits and diseases to provide further clinical context, which highlighted significant genetic overlap with left ventricular mass (correlation 0.81, p = 1.6 × 10^−24^) and the PR interval (correlation 0.26, p = 6.2 × 10^−06^) but did not show strong negative genetic correlations with cardiovascular traits selected for measurement [Fig.S1]. By LDSC, the total observed h2 for IVS.csad was high at 0.068 (SD 0.0106).

Within European ancestry individuals there were two loci which were strongly significant [Table 1, Fig.2A]. A lead variant on chromosome 6 (rs2376620, p_combined_ = 2.00 × 10^−10^, *CDKN1A* intron) which tags a haplotype of linked variants overlapping key open chromatin regions exlusive to ventricular myocardium within the introns of *CDKN1A* [Fig.2C], a canonical regulator of ventricular cardiomyocyte proliferation^41,42^. The other lead variant on chromosome 17 (rs62063281, p_combined_ = 1.31 × 10^−13^) appeared to display strong linkage across a large region of 690,200 base pairs in length which represents a duplication flanked inversion (hg38 chr17:45571611-46261810) across three genes and is common (minor allele frequency 0.18) within in European populations. Using the Akita model of 3D genome folding^21^, we quantified expected changes in chromatin contact frequency caused by the inversion which predicts a severe disturbance in the local chromatin landscape, including loss of contact between the *KANSL1* promoter and regions including the *MAPT* gene body, *KANSL1-AS1*, and several candidate regulatory enhancers predicted by the GeneHancer database [Fig.2B]. A burden-test of structural variants overlapping genes did not reveal further structural variants^19^ [Fig. S2].

**Figure 2.**
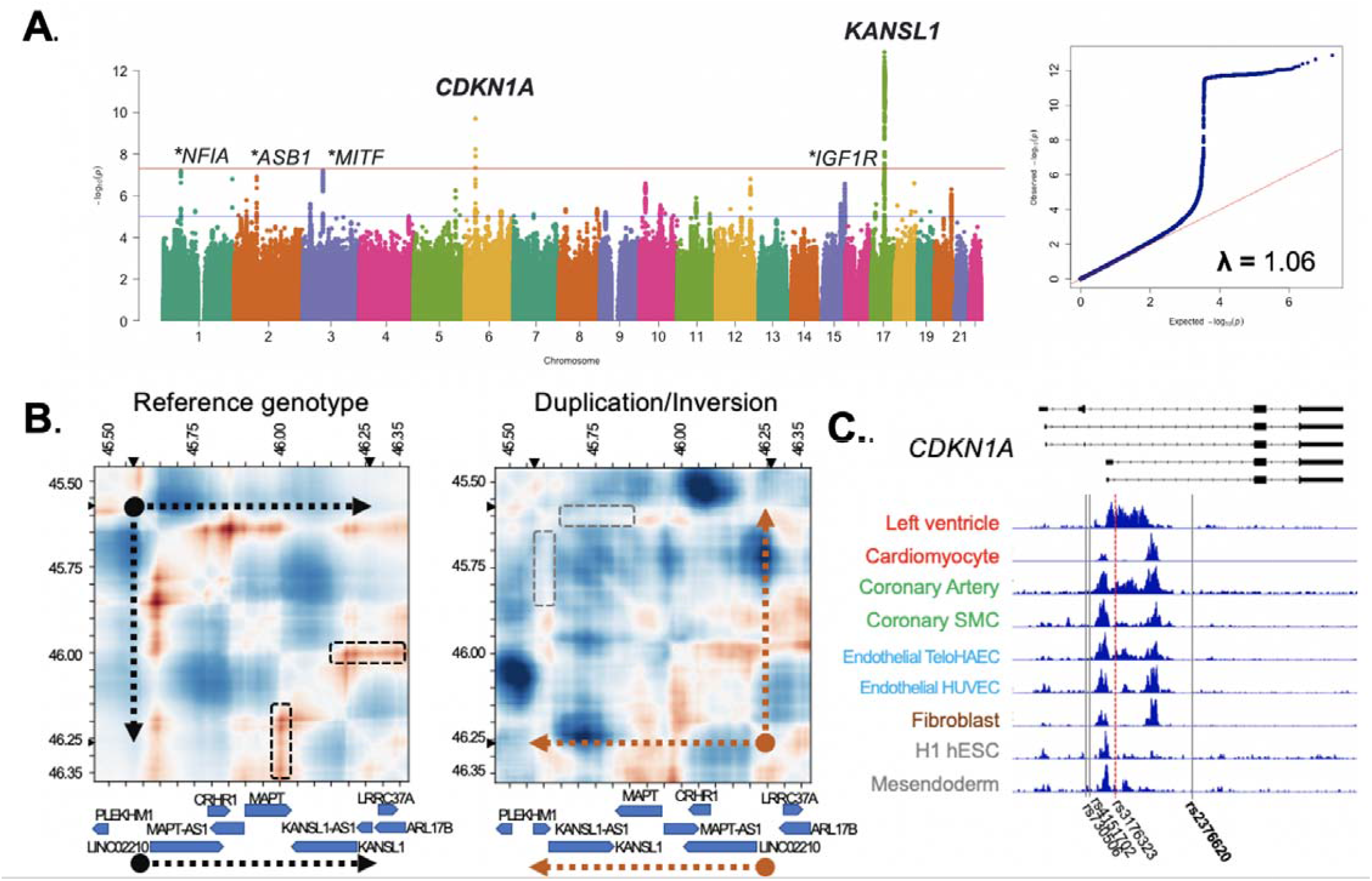
GWAS of IVS.csad identifies two loci meeting standard significance thresholds. **A**. Manhattan plot with two loci at KANSL1 and CDKN1A, along with loci below 5e-08 marked with a star Quantile-quantile plot indicates an absence of systematic inflation. **B**. Two-dimensional plots of the Akita model of genome folding with GRCh38 coordinates on the X and Y axis with relevant genes and strand orientation indicated by blue boxes. Predicted frequency of interaction between regions of DNA is indicated by color, where red(/blue) shows a higher(/lower) frequency of interaction relative to what is expected given linear genomic distance. The reference genotype predicts a strong chromatin interaction (black boxes) between the *KANSL1* promoter and regions including the *MAPT* gene body and *KANSL1-AS1*. The common duplication/inversion 17:45571611-46261810 (orientation indicated by the orange arrow) is predicted to completely disrupt the interaction (corresponding locations within the inversion indicated by the grey boxes). The lead variant rs62063281 is an eQTL for multiple genes across the locus and a strong splicing QTL for *MAPT* and *KANSL1* (−1.5 normalized expression, p=1.7e-88) within the left ventricle [Table S3]. **C**. Plot of tracks showing output of the activity-by-contact (ABC) which identifies *CDKN1A* as the gene target for the lead variant rs2376620, which is the marker for a haplotype inclusive of three other variants localized in open chromatin enhancer regions present most prominently in the left ventricle. Displayed for contrast are DNase-seq for cardiomyocyte, human umbilical vein endothelial cell (HUVEC), fibroblast of fetal lung, H1 human embryonic stem cells (hESCs), and H1-derived mesendodermal precursors; or ATAC-seq for heart left ventricle (LV), coronary artery, coronary smooth muscle cell (SMC), and telomerase immortalized human aortic endothelial cells (TeloHAEC).

**Table 1.**
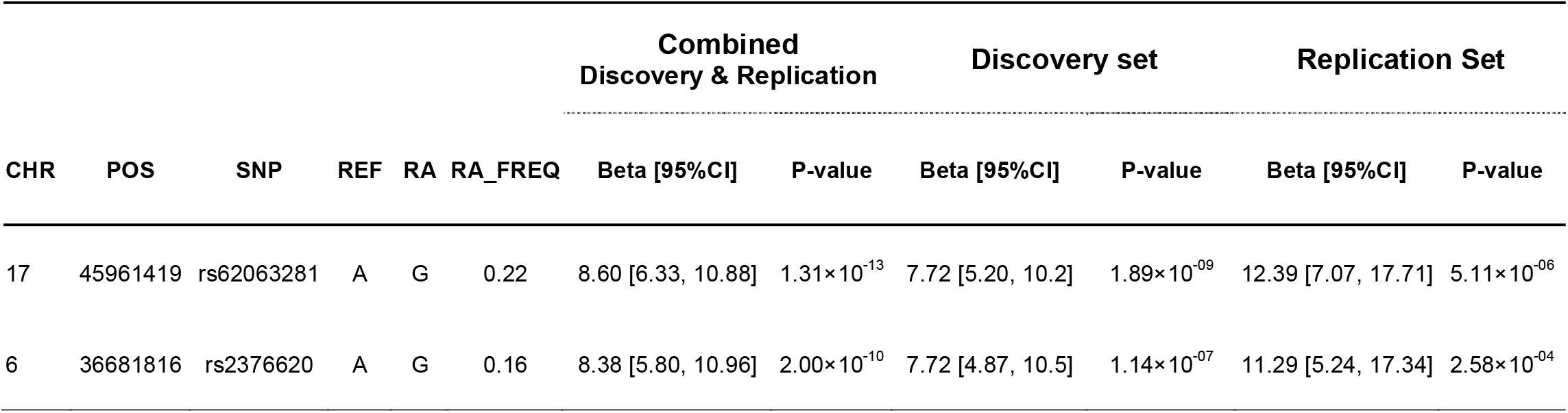
Genome wide associations from measurements of IVS.csad in European ancestry individuals. RA: risk allele, FREQ: frequency

In the European-only GWAS analysis, a number of additional loci did not meet standard genome-wide significance but had strong pre-existing evidence for involvement in cardiovascular biology [Table S4]. Following the European-only analysis, we performed analyses within three additional ethnic strata (South Asian, East Asian, and African/Afro-Caribbean) and a trans-ethnic meta-analysis which confirmed the findings in *CDKN1A* across population strata and identified additional variants on chromosome 3 (rs62253176, p=2.3 × 10^−08^, *MITF* intron) and chromosome 1 (rs2092867, p=4.4 × 10^−08^, *NFIA* intron) which were strongly suggestive but were not meet standard significance threshold in the European only analysis [Figs.S3 & S4]. Additional variants of interest on chromosome 2 (rs1368960, p_combined_ = 1.3 × 10^−07^, *ASB1* intron) and chromosome 15 (rs11633294, p_combined_ = 2.7 × 10^−07^, *IGF1R* intron) displayed clear genomic consequences [Fig.S5] but were not genome-wide significant in European or Trans-ethnic meta-analyses. Additionally we performed both rare variant analyses (MAF 0.01 to 0.001) and gene-burden testing which were unrevealing of genetic associations meeting standard pre-specified significance thresholds [Figs.S6 & S7].

Common genetic variation is known to underpin susceptibility to a variety of cardiovascular diseases, including those determined by extremes in anatomical size.^29,30,31^ We sought to relate the genetic determinants governing normal variation in the IVS to two rare forms of structural heart disease manifesting as either a small or insufficient interventricular septum (VSD) or as a large interventricular septum (HCM). Using an instrumental variable of 22 variants derived from the IVS.csad GWAS [Table S4], we performed two-sample Mendelian Randomization for VSD (191 VSD cases and 5,159 controls) and HCM (2,780 HCM cases and 47,486 controls) as outcome phenotypes. We observed evidence suggesting a causal effect where inheritance of a smaller IVS.csad corresponded to an increase in risk for VSD (beta coefficient: 2.06 log odds ratio (OR) VSD per decrease in SD of IVS.csad, standard error (SE) 0.75, p_IVW_ = 0.006) while inheritance of a larger IVS.csad corresponded to an increase in risk for HCM (beta coefficient: 2.45 log OR HCM per increase in SD of IVS.csad, SE: 0.48, p_IVW_ = 2.8e-7) [Figure 3].

**Figure 3.**
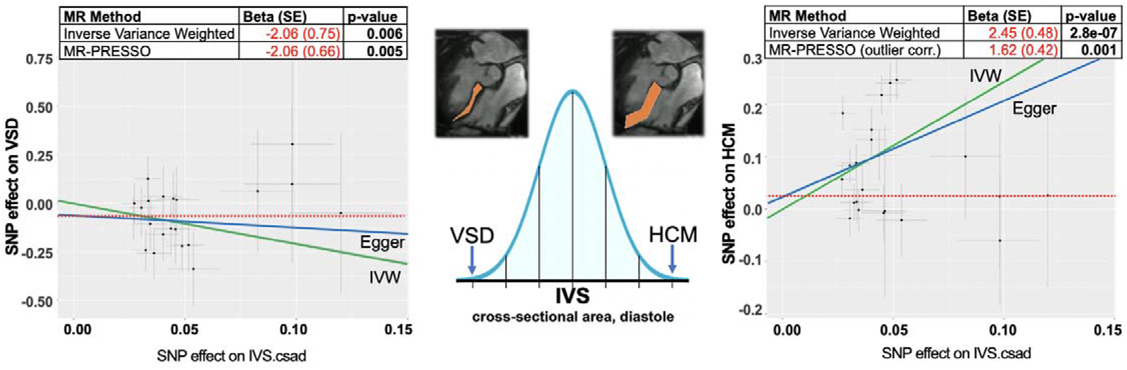
Mendelian Randomization suggests that inheritance of increased IVS.csad is causal for HCM while inheritance of decreased IVS.csad is causal for VSD. Twenty two loci were selected for use as instrumental variables for interventricular septum, cross-sectional area as the exposure for two-sample Mendelian Randomization analysis of ventricular septal defects (VSD) and hypertrophic cardiomyopathy (HCM). Regression lines for the MR-Egger and Inverse Variants Weighted (IVW) methods are disp ayed in blue and green respectively, with the intercept from the MR-Egger method in dotted red for both graphs. The causal relationships of IVS.csad and risks for VSD and HCM are not driven by a single variant when analyzed individually or in a “leave-one out” sensitivity analysis (Figure S6).

When evaluating IVS.csad as the exposure and VSD as the outcome, sensitivity analyses suggested the observed causal effect was uniform across the variants included in the instrumental variable (Q value = 15.8, df=20, p-value=0.73). The MR-Egger test of horizontal pleiotropy was insignificant with broad standard errors around the slope and intercept (beta=-0.62, SE=2.43; intercept=-0.06). The median weighted method, which provides a causal estimate assuming at least 50% of the weight comes from valid instrumental variables, was consistent with the IVW result (beta=-2.40; se=1.05; p=0.02). The MR-PRESSO global test for outliers indicates there was no evidence of horizontal pleiotropy for the outcome of VSD (p=0.78).

Sensitivity analyses evaluating IVS.csad as the exposure and HCM as the outcome suggested there was evidence of heterogeneity within the instrument (Q value = 84.2, df=20, p-value=7.58E-10, i2 = 0.76) and the MR-PRESSO global test for outliers indicated there was horizontal pleiotropy (p = 0.001). The unadjusted MR-PRESSO result (beta coefficient: 2.45, SE: 0.48, p_MR-PRESSO_ = 4.36E-05) is similar is magnitude to the IVW result, but considering the observed horizontal pleiotropy could be artificially inflated. Performing the outlier-corrected MR-PRESSO test (beta coefficient: 1.62 log OR HCM per increase in SD of IVS.csad, SE: 0.42, p_MR-PRESSO_ = 0.001) shows an effect size comparable to the median (weighted) results (1.47 log OR HCM per increase in SD of IVS.csad, SE 0.47, p_median-weighted_ =0.002). The estimate of effect size derived from the MR-Egger test is internally consistent (beta=1.82 log OR HCM per increase in SD of IVS.csad, SE=1.67, p=0.29, intercept=-0.03).

Because left-ventricular mass and IVS thickness are known to increase in the setting of chronic hypertension^10^, a phenomenon that we observed in our clinical correlates of automated measures of IVS.csad [Fig.1B], we performed an additional sensitivity analysis to rule out confounding for the causal effect observed between IVS.csad and HCM. For the outcome of HCM we repeated the two-sample MR for IVS.csad conditioned upon diastolic blood pressure, which did not significantly change the positive and statistically significant causal estimate for IVS.csad (beta 3.08 log OR HCM per increase in SD of IVS.csad, SE 0.48, p_IVW_ = 1.05e-05) [Table S5].

## DISCUSSION

Here we report upon the genetic architecture of the cross-section of the interventricular septum derived from a standard view in cardiac imaging. The measured cross-sectional area of the IVS in diastole is correlated with genetic sex and body surface area along with a clinically accepted measure of the interventricular septal thickness^43^ and genetically correlated with left ventricular mass suggesting that our 2-dimensional automated measure of IVS.csad is a good proxy for the mass of this portion of cardiac anatomy. Genetic associations for IVS.csad are centered upon genetic loci with previously established roles in cardiac development and mendelian forms of cardiac malformations. Additionally, we show a causal relationship between inheritance of a larger interventricular septum with risk of HCM and a smaller interventricular septum with risk of ventricular septal defects.

Haploinsufficiency or truncating mutations in *KANSL1* are causal for Koolen-de Vries syndrome which includes ventricular septal defects amongst a variety of other phenotypes^44,45^. We found that a variant associated with IVS.csad, rs62063281, tags a common inversion on chromosome 17, referred to as CPX_17_4670 on the gnomAD CNV browser^46^, that encompasses *KANSL1* and *KANSL1-AS1* along with four other protein coding genes, and was associated with an increase in IVS.csad (beta= 8.60 mm^2^, p=1.31e-13). The variant rs62063281 which tags the inversion appears to be a strong tissue-specific splice QTL (Normalized expression −1.5, p = 1.7e-88) for *KANSL1*, In addition to disruption of the interaction of the *KANSL1* promoter and predicted regulatory elements, genes that contact *KANSL1-AS1* (including *ARL17B, LRRC37A, NSFP1, LRRC37A2, NSF*) in the reference configuration are also expected to lose this interaction as a result of the common inversion, which potentially may additionally result in disrupted expression of these other genes. As a group histone, modifying genes including the WDR5-MLL1 complex of which KANSL1 is a component are well-recognized to be involved in cardiac development and diverse forms of congenital heart disease^5,47^. Together these data suggest that common alterations to dosage and transcription of *KANSL1* impact the development and growth of the interventricular septum.

The *CDKN1A* locus has been specifically identified as a target of HIF-1a within the developing ventricular septum^48^ and has been experimentally described as a key mediator of the hypertrophic response within postnatal life^41,49^. The variant rs2376620 sits within an intron of *CDKN1A*, a locus well known to play a role in cardiomyocyte growth^42^ recently identified in heart failure^29^. Together these data suggest a specific role for *CDKN1A* in normal development of the interventricular septum along with physiological function throughout postnatal life. Among the other identified in the trans-ethnic analysis, the variant rs62253176 occurs within an intron of the transcription factor *MITF*, an important mediator of beta-adrenergic induced hypertrophy via the renin-angiotensin system expressed in cardiomyocytes^50^. Importantly, *MITF* binds the promotor of canonical cardiac development transcription factor *GATA4* and knockdowns in human embryonic stem cell derived cardiomyocytes specifically reduce the expression of genes in the sarcomere^51,52^. The role of *NFIA* in cardiac development and function is less well defined though it is associated with a variety of electrocardiographic traits^53,54^ and the variant rs2092867 occurs on a haplotype associated with a recently reported novel trait related to the presence of cardiac trabeculae derived from the same UK Biobank imaging dataset^55^.

It is well established that rare causal variants confer large effects and produce extreme phenotypes. However, many individuals possessing an extreme phenotype do not yield such causal variants when subject to clinical genetic testing^56^. Instead, there is increasing evidence that such extreme phenotypes may also be attributable to the aggregate burden of common variants associated with the disease or a causal trait^57^. As such, extremes in polygenic risk may explain variability in cardiovascular anatomy and represent an important disease risk factor for a variety of different diseases. For instance, we have previously shown that polygenic risk scores influence ascending aorta size (PRS_Aorta_); increases in PRS_Aorta_ are associated with risk of developing thoracic aortic aneurysm and dissection^58^, and decreases in PRS_Aorta_ are associated with left ventricular outflow tract congenital heart disease^59^.

Using two-sample Mendelian randomization, we show that inheritance of a smaller IVS.csad results in an increase in ventricular septal defect risk, and conversely, inheritance of a larger IVS.csad confers increased risk in HCM. Identifying risk of ventricular septal defects is increased through the inheritance of a smaller interventricular septum, as measured by IVS.csad, supports the need for systematic inquiry into the relationship between normal variation in the size and shape of cardiovascular anatomy and congenital heart disease, for which the majority of genetic risk remains unexplained. For HCM, rare pathogenic variants within core sarcomere genes account for 40% of HCM cases, but with unexplained phenotypic heterogeneity. Recent data suggest common genetic variants also underpin HCM risk^36^. The analyses presented here suggest that a proportion of HCM risk is attributable to normal variation in the size and mass of the interventricular septum as measured by IVS.csad [Fig.S2].

There are limitations to each aspect of our study. While we have created an automated measure of the interventricular septum which is simple in conception and shown correlation with a variety of traits, the measure lacks phenotypic context provided by many years of use in standard clinical imaging. The cross-sectional area of the interventricular septum is a reductive two-dimensional measure of a complex three-dimensional structure which changes dynamically over the course of the cardiac cycle^60^. More complex measures of the septum are likely to provide nuanced correlations with clinical phenotypes, patient outcomes, as well as substrate for discovery of novel genetic determinants of structure and function^55^. Our analyses were limited by the small proportion of individuals of African/Afro-Caribbean, South and East Asian descent, which reduced the power to detect genetic signal within these groups for this trait and underscores the urgent need to diversify genetic studies of all human diseases and traits^53^. While the inclusion of structural variation in our association studies is novel and our finding is supported by the association of *KANSL1* with Mendelian diasease and mouse models of cardiac development, there are no cohorts available for external confirmation which include both CNV calls and readily available bespoke measures of cardiac anatomy. Although our Mendelian randomization analyses yielded clear and consistent results relating the heritable architecture of septal architecture to rare cardiovascular diseases, they are limited in scope by the relatively underpowered outcome GWAS for VSD and the clinical utility of these findings require further contextualization in larger cohorts.

Overall, the data and analyses presented here illustrate the power of combining genetics with phenotyping of cardiac imaging accelerated by machine learning to identify new loci related to cardiovascular development and pathology. The study of cardiovascular anatomy measured as a continuous trait offers the potential to disentangle complex risk factors and reveal previously unrecognized heritability for rare forms of structural heart disease. While the risk is primarily thought to be inherited, the genetic architecture of congenital heart disease has been difficult to establish. In addition other evidence^59^, the relationship between a small IVS to increased risk of VSD may suggest a that a proportion of genetic risk for other forms of congenital heart disease can be localized to heritable extremes in size of cardiovascular anatomy which arise primarily from common genetic variation.

## Data Availability

All data are available from the UK Biobank organization.

## ACKNOWLEDGEMENTS

The authors wish to thank Dan Bernstein, James Pirruccello, and Euan Ashley for helpful commentary on the manuscript.

## FUNDING

NIH NHGRI R35HG011324 (to JME), Gordon and Betty Moore and BASE Initiative at the Lucile Packard Children’s Hospital at Stanford University (to JME), American Heart Association / Children’s Heart Foundation Congenital Heart Defect Research Award (to DA), Stanford Maternal and Child Health Research Institute (to DA). JRP has been supported by the Stanford University Department of Pediatrics, National Heart Lung and Blood Institute (R00 HL130523), and Chan-Zuckerberg Biohub.

## Notes

### Competing Interest Statement

Dr. Priest is an employee and shareholder of Tenaya Therapeutics which has publicly disclosed development programs in Hypertrophic Cardiomyopathy.
Dr. Watkins is a paid consultant to BioMarin
Dr. Harper is an employee of AstraZeneca

### Funding Statement

No authors or institutions received payment or services from a third party for any aspect of the submitted work.
JRP has been supported by the National Heart Lung and Blood Institute (R00 HL130523) and Chan-Zuckerberg Biohub.

