## Supplemental Tables for "Genetic determinants of interventricular septal anatomy and the risk of ventricular septal defects and hypertrophic cardiomyopathy"

| **Phenotypes** | **EUR combined** | **EUR**  **discovery** | **EUR**  **replication** | **AFR** | **EAS** | **SAS** |
| --- | --- | --- | --- | --- | --- | --- |
|  | **sample size** | **sample size** | **sample size** | **sample size** | **sample size** | **sample size** |
| **IVS.csad** | 26,844 | 21,945 | 4,899 | 214 | 84 | 365 |

**Table S1. Sample sizes of population strata combining genetic data and automated measures of interventricular septum cross-sectional area at diastole.**

**Table S2. Linear modeling of systematic error of automated measurement of IVS.csad**

| **Characteristic** | **Beta coefficient** | **95% Confidence Interval** | **p-value** |
| --- | --- | --- | --- |
| Body surface area (m^2^) | -0.1582 | -0.3578 to 0.0415 | 0.118 |
| Genetic sex (Female) | -0.0008 | -0.09 to 0.07 | 0.830 |
| Age (years) | -0.0002 | -0.007 to 0.002 | 0.330 |

Percent error ((automated ML measurement – human measurement) / human measurement) for 50 randomly selected individuals is not predicted by Body surface area, Genetic sex, or Age at enrollment in standard linear modeling. Adjusted R-squared = 0.05, p=0.1429. Mean error across all 50 individuals was -3.7% or -29 mm^2^.

| **Gene Symbol** | **SNP Id** | **P-Value** | **Normalized Exression** | **Tissue** |
| --- | --- | --- | --- | --- |
| *ARL17A* | rs62063281 | 2.00E-14 | 0.65 | Heart - Left Ventricle |
| *DND1P1* | rs62063281 | 3.30E-64 | 1.3 | Heart - Left Ventricle |
| *KANSL1-AS1* | rs62063281 | 4.80E-80 | 1.2 | Heart - Left Ventricle |
| *LINC02210* | rs62063281 | 4.90E-89 | 1.1 | Heart - Left Ventricle |
| *LRRC37A* | rs62063281 | 1.30E-09 | 0.5 | Heart - Left Ventricle |
| *LRRC37A2* | rs62063281 | 1.20E-56 | 1 | Heart - Left Ventricle |
| *LRRC37A4P* | rs62063281 | 1.40E-68 | -1.1 | Heart - Left Ventricle |
| *MAPK8IP1P2* | rs62063281 | 4.10E-70 | 1.2 | Heart - Left Ventricle |
| *MAPT* | rs62063281 | 3.00E-16 | -0.48 | Heart - Left Ventricle |
| *RP11-259G18.1* | rs62063281 | 1.70E-12 | 0.57 | Heart - Left Ventricle |
| *RP11-259G18.3* | rs62063281 | 6.80E-76 | 1.2 | Heart - Left Ventricle |
| *RP11-707O23.1* | rs62063281 | 6.50E-39 | 1 | Heart - Left Ventricle |
| *WNT3* | rs62063281 | 2.90E-09 | 0.48 | Heart - Left Ventricle |
| **Gene Symbol** | **SNP Id** | **P-Value** | **Normalized Exression** | **Tissue** |
| *KANSL1* | rs62063281 | 1.70E-88 | -1.5 | Heart - Left Ventricle |
| *MAPT* | rs62063281 | 0.0000043 | 0.43 | Heart - Left Ventricle |

**Table S3. Left ventricle-specific eQTL and sQTL data for the lead variant on Chromosome 17 which tags a common inversion.** Data extracted from GTEx version 8.

| **SNP** |  | **Manual annotation** | **Additional** | **IVS.csad** | **IVS.csad** | **HCM** | **VSD** |
| --- | --- | --- | --- | --- | --- | --- | --- |
|  | **Chr:Loc:Ref:Alt** |  | **Information** | **Beta [SE]** | **P value** | **P value** | **P value** |
| rs182036013 | chr10:79857619:A:C | intergenic |  | 16.3 [3.6] | 4.54E-06 | 0.86 | 0.81 |
| rs58864905 | chr8:23763707:T:G | AC012574.1  lincRNA intron |  | 5.6 [1.2] | 4.48E-06 | 0.74 | 0.93 |
| rs4307773 | chr12:50750649:T:C | intergenic |  | 4.5 [1] | 4.39E-06 | 0.07 | 0.45 |
| rs1057410 | chr10:73814321:G:A | 3’ UTR *CAMK2G* |  | 6.7 [1.4] | 2.94E-06 | 5.09E-04 | 0.82 |
| rs34397544 | chr17:16507722:A:G | intergenic |  | -9 [1.9] | 2.59E-06 | 0.76 | 0.08 |
| rs6768396 | chr3:25380216:A:G | Intron *RARB* | Syndromic form of VSD (OMIM 615524) | 6 [1.3] | 2.53E-06 | 0.42 | 0.05 |
| rs8039472 | chr15:84818413:A:G | Intron *ALPK3* | Mendelian form of HCM (617608) | 4.5 [1] | 2.50E-06 | 1.84E-08 | 0.98 |
| rs1346 | chr11:65569780:A:T | Non-coding RNA exon |  | 5.7 [1.2] | 2.23E-06 | 0.96 | 0.44 |
| rs114510001 | chr2:43289949:C:T | Intron *THADA* |  | -7.7 [1.6] | 1.63E-06 | 0.98 | 0.92 |
| rs76833326 | chr20:59185122:C:T | intergenic |  | 7.6 [1.5] | 8.55E-07 | 0.89 | 0.46 |
| rs72831141 | chr5:148156177:G:A | intergenic |  | -13.8 [2.8] | 5.67E-07 | 0.39 | 0.84 |
| rs11633294 | chr15:98731779:C:A | Intron *IGF1R* | Signaling pathway in cardiac hypertrophy & development (PMID **30909860)** | -5.4 [1] | 2.69E-07 | 0.73 | 0.02 |
| rs55716025 | chr10:21114754:G:A | Intron *NEBL* | Component of sarcomere  (PMID **26321576)** | -7.3 [1.4] | 2.62E-07 | 0.03 | 0.36 |
| rs145567583 | chr18:67621286:C:T | Intron ncRNA RP11-638L3.1 |  | 20 [3.9] | 2.55E-07 | 0.88 | 0.9 |
| rs6547225 | chr2:79176812:T:G | Intron ncRNA AC011754.1 |  | 5.1 [1] | 1.81E-07 | 0.6 | 0.52 |
| rs71648698 | chr1:242231277:C:T | Intron *PLD5* |  | -16.4 [3.1] | 1.58E-07 | 0.63 | 0.48 |
| rs9795600 | chr12:121486529:G:T | Intron *KDM2B* | Mouse knockouts with defects in cardiac development (PMID 25848754) | 5.1 [1] | 1.57E-07 | 0.01 | 0.82 |
| rs2207792 | chr1:61429037:G:A | Intron *NFIA* | Discussed in main text | 5.6 [1] | 6.41E-08 | 0.01 | 0.26 |
| rs7623486 | chr3:69859943:A:G | Intron *MITF* | Discussed in main text | 6.7 [1.2] | 6.17E-08 | 1.22E-03 | 0.24 |
| rs6937605 | chr6:36692155:C:T | Intron CDKN1A | Discussed in main text | 7.5 [1.3] | 5.97E-09 | 9.02E-09 | 0.87 |
| rs55938136 | chr17:45720994:A:G | Inversion KANSL1 | Discussed in main text | 8.1 [1.2] | 1.99E-12 | 2.50E-10 | 0.1 |
| rs62063281 | chr17:45961419:A:G | Intron CRHR1 | Component of large inversion | 8.6 [1.2] | 1.31E-13 | 1.57E-10 | 0.11 |

**Table S4. Variants related to IVS.csad selected for use as instrumental variables in Mendelian Randomization analyses with summary statistics for the primary GWAS, and p-values each of the two outcome GWAS (Cordell et. al, Harper et. al.).** Note that Beta coefficients are presented here in mm^2^ for consistency with Table 1, while the Mendelian randomization analyses incorporated standardized beta coefficients. Annotations performed manually examing the UCSC Genome Browser and GTex v8.

| **Coefficient** | **beta** | **se** | **t-value** | **p-value** |
| --- | --- | --- | --- | --- |
| IVS.csad | 3.08 | 0.48 | 6.473 | 1.05e-05 |
| DBP | -0.304 | 0.135 | -2.257 | 0.0394 |

**Table S5. Sensitivity analysis for IVS.csad 🡪 HCM conditioned upon diastolic blood pressure.** After accounting for diastolic blood pressure, the instrumental variable for IVS.csad displays a causal estimate (beta 3.08, SE 0.48, p_IVW_ = 1.05e-05) which is even slightly larger than the primary causal estimate presented in the main text (Fig.3). Results are strongly suggestive that the positive causal estimate detected does not arise from reverse-causality or pleiotropy within the component SNPs included in the instrumental variable.
